# Dysregulated smooth muscle cell proliferation and gene expression underlie ACTA2 variant-associated aortopathy

**DOI:** 10.1101/2025.08.04.25331815

**Authors:** Mark E. Pepin, Richard Mitchell, Rajat M. Gupta

## Abstract

Pathogenic variants affecting alpha-2 smooth muscle actin (ACTA2) account for approximately 20% of non-syndromic familial thoracic aortic aneurysms (TAA) and confer a high risk of dissection; however, the cell-type-specific transcriptional mechanisms underlying□ACTA2-associated TAA remain poorly defined, particularly for variants of uncertain significance. In this study, we investigated the transcriptional and cellular effects of a novel ACTA2 p.Met49Thr mutation identified in a young male in his early twenties who developed a dissected ascending aortic aneurysm without traditional risk factors. Using the clinically archived formalin-fixed paraffin-embedded (FFPE) aortic tissue, we isolated intact nuclei and performed single-nucleus RNA sequencing (snRNA-seq) to generate 17,938 transcriptomes. Relative to non-genetic hypertensive TAA control, The ACTA2-p.Met49Thr sample displayed marked expansion of vascular smooth muscle cells (VSMCs) relative to TAA (70.6% vs. 39.7%), accompanied by upregulation of proliferation-associated transcripts including FOSB, FOS, JUN, and DEPP1. Lineage tracing via trajectory analysis revealed a transcriptional progression from quiescent to pro-proliferative VSMC states enriching for human loci associated with aortic strain and diameter. Histological evaluation corroborated these findings, demonstrating medial hypercellularity, elastic fiber fragmentation, and adventitial fibrosis enriched within the ACTA2-p.Met49Thr specimen. Taken together, these findings implicate a novel pathogenic ACTA2 variant that drives transcriptional reprogramming and proliferative VSMC remodeling, supporting that ACTA2-associated aortopathy progression occurs via functional cell state transitions. Additionally, this work demonstrates the feasibility of FFPE-compatible snRNA-seq as a useful tool for clinical variant annotation.

## Introduction

Alpha 2 Smooth Muscle Actin (ACTA2) mutations are implicated in nearly 20% of non-syndromic familial thoracic aortic aneurysms (TAA),^1^ conferring a high lifetime risk of dissection or aneurysm formation.^2^ Despite its causal role, the specific cell-type signaling mechanisms driving ACTA2-mutant TAA pathogenesis remain poorly understood. In this report, we expand on recently published evidence that specific ACTA2 mutations affect transcriptional regulation in aortic smooth muscle to promote a pro-proliferative vascular smooth muscle cell phenotype. We apply a novel single-nucleus RNA-sequencing (snRNA-seq) method compatible with clinical formalin-fixed vascular biospecimens to identify the transcriptional changes that occur in vascular cells from an aortopathy patient who was found to harbor a previously uncharacterized ACTA2 mutation. We identify the expansion of a pro-proliferative VSMC subpopulation with widespread transcriptional cell state shifts. These findings validate the altered chromatin remodeling changes recently found for R179C ACTA2 mutations associated with a more pervasive smooth muscle dysfunction syndrome seen in children.^3^ More broadly, we used snRNA-seq technology to streamline the clinical implication of a novel variant that validates preclinical evidence that ACTA2 mutations drive disease through pathologic cell state shifts among aortic VSMCs.

## Results

### Single-nucleus RNA sequencing (snRNA-Seq) of ACTA2-p.M49T aortic tissue reveals vascular smooth muscle cell expansion and distinct transcriptional clusters

We encountered a young man in his early twenties with no knowledge of pre-existing medical conditions who developed a 5.3-cm Stanford Type-A ascending aortic aneurysm which had dissected into his right brachiocephalic, left subclavian and left common carotid arteries after developing recurrent episodes of right-sided neck and arm paresthesia, pain, arm numbness, lightheadedness, blurred vision, and headache (**Fig. 1A**). He had no prior medical history consistent with smooth muscle dysfunction syndrome. Vascular imaging did not identify other lesions including in the renal, coronary, or intracranial arteries. Following an emergent ascending aortic and zone 2 aortic arch and valve-sparing aortic root replacement, he underwent genetic screening which identified a heterozygous single nucleotide polymorphism (SNP) in exon 3, c.146T>C (p.Met49Thr) of the ACTA2 gene (rs869025352). This mutation results in the substitution of methionine, a neutral and non-polar amino acid, with threonine, a neutral and polar amino acid, at codon 49 of exon 3 (**Fig. 1B**).

**Figure 1:**
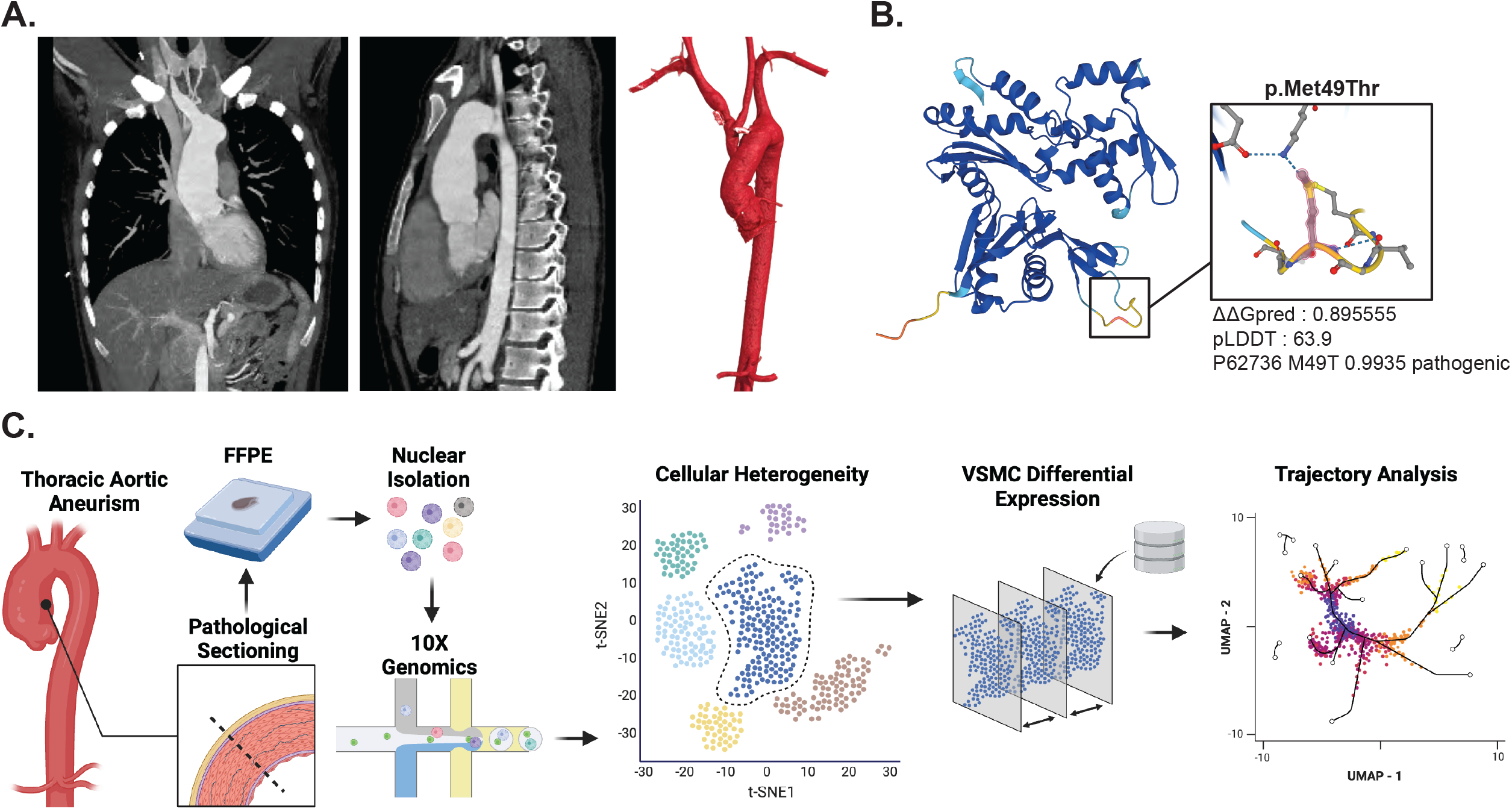
Subject characterization and analytic workflow p.Met49Thr mutation. **(A)** Computed tomography scan using arterial contrast to demonstrate a 5.3-cm Stanford Type A ascending aortic aneurysm with dissection flap extending into the right brachiocephalic artery, along with a 3-dimensional reconstruction of the surgical repair. **(B)** Predicted protein structure of Alpha smooth muscle actin (ACTA2) based on AlphaFold® prediction, highlighting methionine residue at position 49. **(C)** Visual representation of the analysis of nuclei extracted from formalin-fixed paraffin-embedded (FFPE) tissue used for single-nuclear RNA-sequencing analysis.

Given a lack of fresh tissue needed to functionally characterize this variant in our patient, we tailored a protocol to extract nuclei from formalin-fixed and paraffin-embedded (FFPE) aortic tissue collected during his aortic repair for clinical histology. To accomplish nuclei dissociation and isolation from FFPE, we adapted a previously described protocol using 50µm FFPE scrolls dissociated with the Miltenyi Biotech FFPE Tissue Dissociation Kit, filtered through a 20-μm strainer to remove debris, stained with DAPI for viability assessment via fluorescence microscopy, and analyzed for RNA quality using the TapeStation platform.^7^ We loaded ∼20k intact nuclei onto a Chromium Q chip (PN-1000422, 10X Genomics) to perform single-nucleus RNA sequencing (snRNA-seq) to explore ACTA2-p.M49T variant at single-nucleus resolution (**Fig. 1C**), which we provide as a public resource: https://markpepin.shinyapps.io/ACTA2_pMet49Thr_snRNA_Seq/. To identify the cell type-specific transcriptional shifts that accompany this ACTA2-associated aortopathy, we integrated the snRNA-seq datasets of the ACTA2-p.M49T subject with that of aortic tissue from a patient with acquired TAA attributed to poorly-controlled hypertension and without evidence of familial aortopathy or syndromic features (TAA-NG) (**Fig. 2A**). Unsupervised clustering via UMAP identified 11 distinct clusters (**Fig. 2B**), which we found to correspond with two distinct vascular smooth muscle (VSMC_1/2), Fibromyocyte, Fibroblast, B Lymphocyte (B Lymph), T Lymphocyte (T Lymph), Natural Killer T (NKT), Macrophage, Dendritic, and Endothelial (DC) cells (**Fig. 2C-D**). Quantification of relative cell-type abundance revealed a higher proportion of VSMCs (**Fig. 2E**), wherein the ACTA2-p.M49T mutant sample contained 70.6% VSMCs (12,385/17,532 nuclei) compared to 39.7% VSMCs (4,646/11,716 nuclei) within the TAA-NG sample. Differential expression between ACTA2-p.M49T and TAA-NG across cell types uncovered a broad array of overlapping – and distinct – differential gene expression (**Fig. 2F**). VSMC subtype 1 (VSMC_1) was found to exhibit the highest number of differentially-expressed genes (DEGs) relative to DEGs among other cell types. Among these, the most differentially expressed in ACTA2-p.M49T revealed FOSB, SORBS1, C11orf96, ACTC1, SOCS3, and DEPP1 as broadly up-regulated across cell types (**Fig. 2G**). Clustered gene-set enrichment analysis identified conserved “extracellular matrix organization” (FDR = 3.0 × 10^−6^, Enrichment = 14/74) and others associated with inflammatory signaling (**Fig. 2H**).

**Figure 2:**
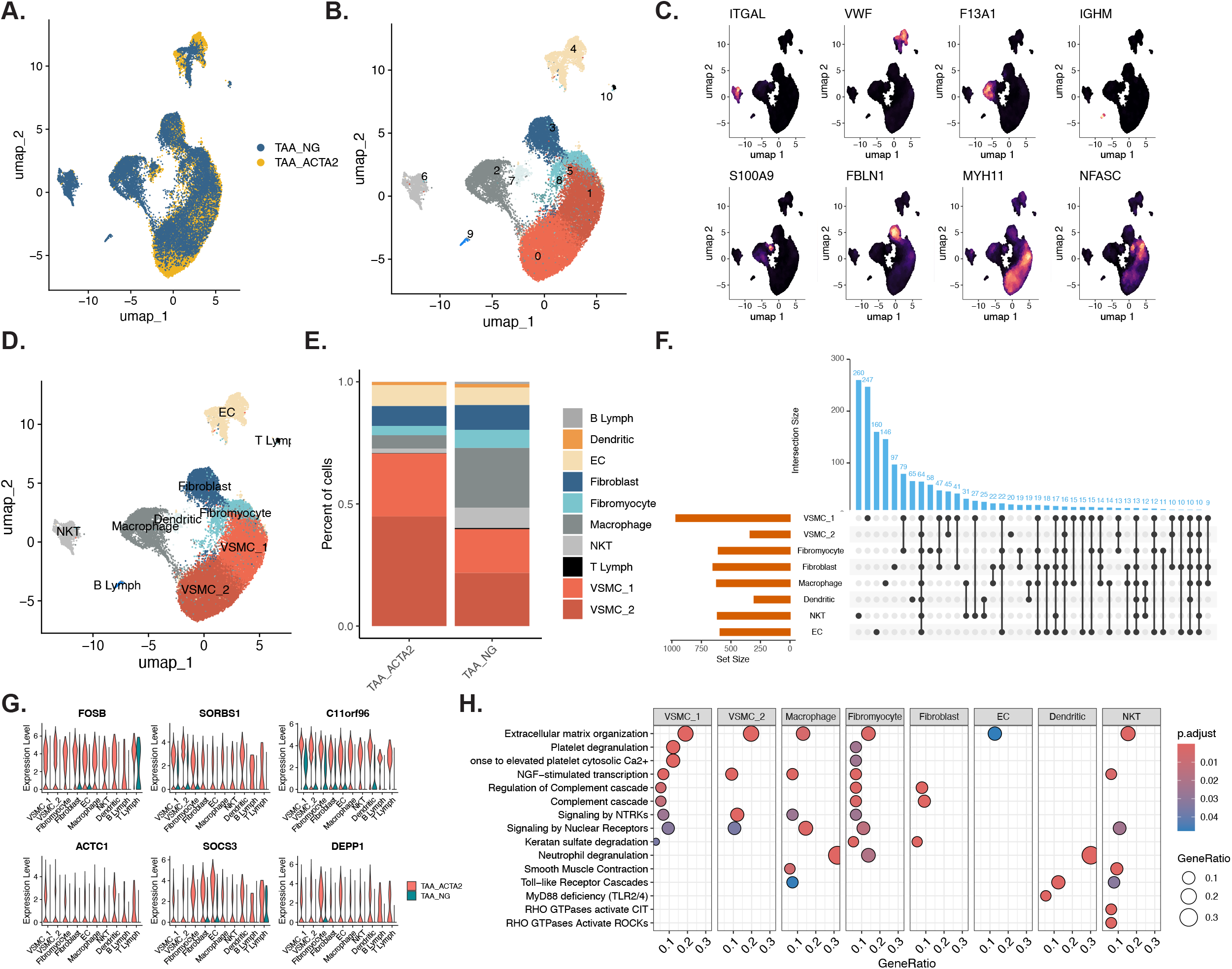
Integrated single-nuclear transcriptomic analysis identifies disproportionate enrichment of vascular smooth muscle cells. **(A)** UMAP-based unsupervised clustering following integration of ACTA2-p.Met49Thr mutation (TAA_ACTA2) and hypertensive aortopathy (TAA_NG). **(B)** UMAP-based clustering at 0.4 resolution, identifying 11 distinct clusters. **(C)** Density plots of gene markers for known cell types, identifying **(D)** Endothelial Cell (EC), T Lymphocyte (T Lymph), Fibroblast, Fibromyocyte, Dendritic, Macrophage, B Lymphocyte (B Lymph), Natural Killer T Lymphocyte (NKT), and two clusters of Vascular Smooth Muscle Cell (VSMC_1/2) populations. **(E)** Proportional bar plot showing relative enrichment of VSMCs in TAA_ACTA2. **(F)** UpSet plot illustrating the intersection of differentially expressed genes (DEGs)* in TAA_ACTA2 relative to TAA_NG across cell types, highlighting shared and unique gene sets within the dataset. **(G)** Violin plot of gene expression profiles of 6 shared DEGs across cell types. **(H)** Clustered gene set enrichment analysis to overlapping pathways across cell types, showing the ratio of DEGs to the number of genes in the pathway (GeneRatio) and BH-corrected p-value (p.adjust) based on the Fisher’s Exact test. *Statistical significance is assumed based on a BH-corrected p-value < 0.05.

### Pathological Cell State Transition of VSMCs in ACTA2-p.M49T Thoracic Aortic Aneurysms

Given the observed expansion of VSMCs in the ACTA2-p.M49T-associated TAA, we next focused on identifying gene expression changes that might underlie this pathological phenotypic shift and potential cellular expansion. Following re-clustering and cellular labelling of the ACTA2-p.M49T variant sample (**Fig. 3A-B**), we performed differential expression analysis comparing the VSMC_1 pool relative to VSMC_2 (**Fig. 3C**). This analysis revealed the most robustly upregulated genes as those associated with cellular proliferation, including proto-oncogenes FosB (FOSB), Fos (FOS), Jun (JUN), zinc finger protein 36 (ZFP36), and DNA damage inducible transcript 4 like (DEPP1). Although these genes displayed increased expression from the VSMC_1 to VSMC_2 clusters, the differential expression was significantly more pronounced among smooth muscle cells harboring the ACTA2-p.M49T mutation compared to those from TAA-NG (**Fig. 3D**). Furthermore, these genes were nearly undetectable in the TAA-NG sample regardless of sub-cluster identity.

**Figure 3:**
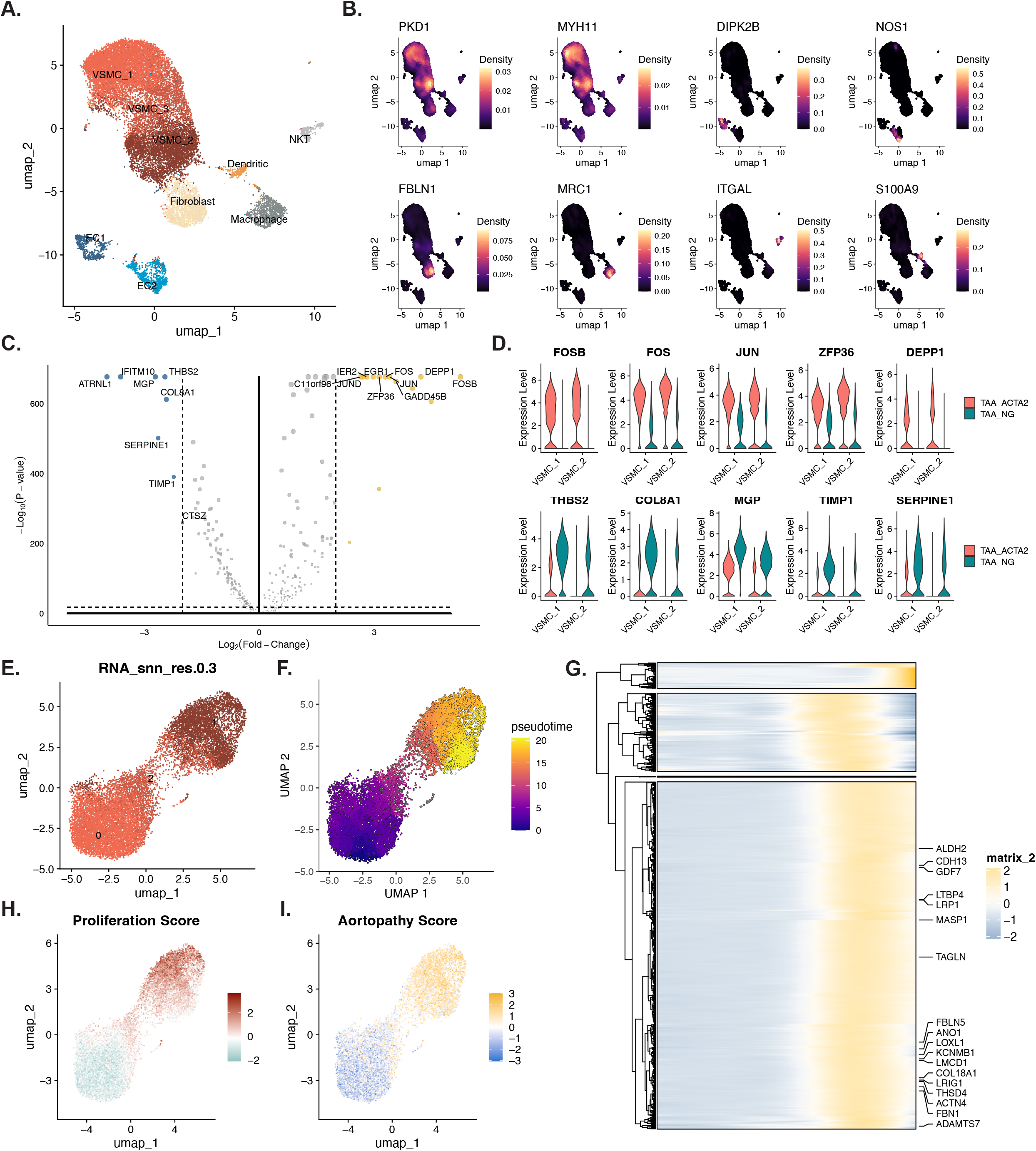
Single-nuclear RNA Sequencing of ACTA2-p.Met49Thr uncovers a pro-proliferative cell state transition within vascular smooth muscle fraction. **(A)** Unsupervised UMAP clustering of ACTA2-p.Met49Thr aorta reproduces canonical cell types, as determined using **(B)** densitometric visualization of gene markers for VSMC (PKD1, MYH11), EC (DIPK3B, NOS1), Fibroblast (FBLN1), Macrophage (MRC1), NKT (ITGAL), and Dendritic (S100A9) cells. **(C)** Volcano plot of all differentially expressed genes in the ACTA2-p.Met49Thr relative to non-genetic (TAA_NG) sample, with **(D)** Violin Plot of top-5 most up-regulated and down-regulated DEGs. **(E)** Sub-grouping of VSMCs identifies 3 distinct clusters at 0.3 UMAP resolution. **(F)** Visualization of “Proliferative score,” representing the scaled sum of all constituent genes within the GO-term GO0008283 as performed Kwartler by et al.^3^ **(G)** Trajectory analysis of VSMCs using proliferative score to identify origin. **(H)** Hierarchical clustering and heatmap visualization of genes significantly altered across pseudotime, showing splined counts. **(I)** UMAP visualization of scaled sum of significantly enriched genes associated with variants linked to aortophathy traits (“Aortopathy score”) using the 2023 GWAS database.

Based on these observations, and in light of prior evidence that ACTA2 mutations promote a pro-proliferative phenotype in murine VSMCs,^7^ we used trajectory analysis of the VSMC population (**Fig. 3E**) to demarcate pathological cell state transitions within the VSMC fraction of ACTA2-p.M49T (**Fig. 3F**). This analysis identified three distinct patterns of gene expression across pseudotime, wherein most genes underwent transcriptional activation between the transition between VSMC_2 and VSMC_1. Gene set enrichment within this trajectory identified a variety of traits linked to aortopathy, including “pulse pressure” (22/735 genes, *P* = 4.1 × 10^−7^), “systolic blood pressure” (23/1073, *P* = 5/4 × 10^−5^), and “diastolic blood pressure” (16/803, *P* = 0.0017).

Building on the observation that ACTA2-p.M49T is associated with VSMC_1-specific upregulation of pro-proliferative genes, we quantified this shift using a composite “proliferation score” to assess transcriptional changes within human VSMC clusters (**Fig. 3H**), as previously reported.^7^ This score, calculated as the z-score of genes associated with the proliferation-related GO term GO:0008283, mirrored the predominant trend of activated genes across pseudotime wherein the VSMC cell state transition exhibited a gradient of pro-proliferative gene activation. Mapping pseudotime differentially expressed genes (DEGs) to disease-associated genetic loci identified via human GWAS revealed a similar transition, with GWAS-enriched DEGs increasingly represented within the VSMC_2 population. (**Fig. 3I**). Specifically, traits enriched within this VSMC subpopulation included “Descending Thoracic Aortic Diameter” (FDR = 0.0029, 10/43 genes), “Descending Aorta Maximum Area (MTAG)” (FDR = 0.007, 7/24 genes), “Descending Aorta Strain” (FDR = .008, 5/11 genes), and “Descending Aorta Distensibility” (FDR = 0.02, 4/8 genes). The common underlying genes responsible for these terms include ALDH2, LTBP4, LRP1, MASP1, TAGLN, ACTN4, FBN1, CDH13, ANO1, LOXL1, KCNMB1, LMCD1, COL18A1, LRIG1, THSD4, and ADAMTS7. Taken together, these preliminary observations support that the ACTA2-p.M49T variant is associated with a disease-relevant shift among VSMCs to promote a pro-proliferative transcriptional phenotype.

### ACTA2-p.Met49Thr is associated with increased aortic elastic tissue fragmentation, adventitial fibrosis, and smooth muscle cellularity in ACTA2-p.M49T

To determine whether the transcriptional cell state transition promoting pro-proliferative expression reflects a morphological shift in aortic composition, we used the same aortic cross-sections for histologic staining (**Fig. 4**). The ACTA2-p.M49T aorta exhibited elastic tissue fragmentation and adventitial fibrosis, occurring notably in the absence of substantial inflammatory cell infiltration. Within the medial layer, there was greater smooth muscle cellularity. By contrast, histologic analysis of the TAA-NG subject demonstrated a substantially lesser degree of elastic tissue fragmentation, minimal adventitial fibrosis, and normal degree medial cellularity. Taken together, these histologic differences support that the pro-proliferative transcriptional signature reflects distinct pathologic characteristics associated with the ACTA2-p.M49T variant relative to non-genetic aortopathy.

**Figure 4:**
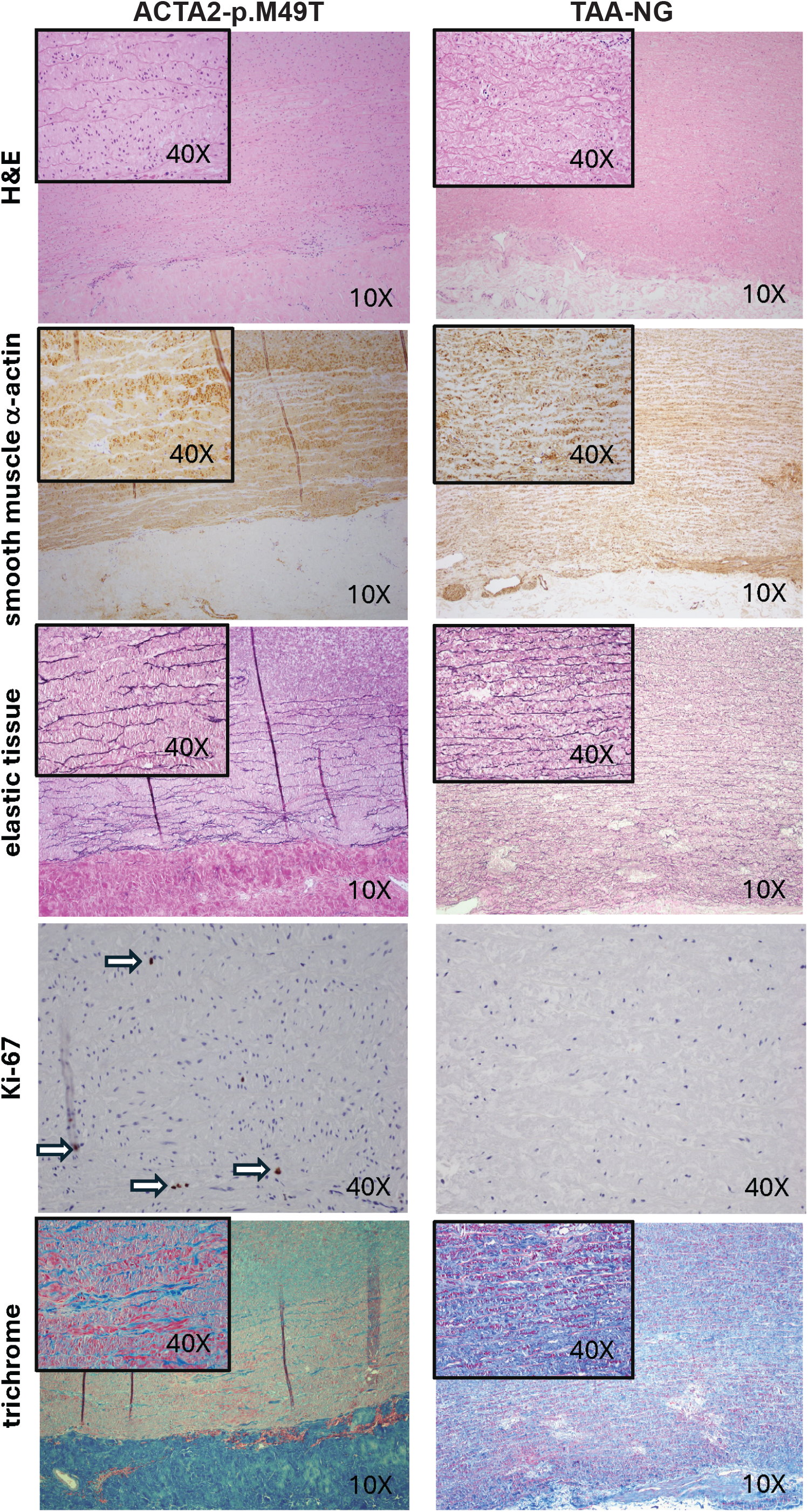
Histologic analysis. Stains and magnification are indicated in each panel. The first column shows staining from the index patient harboring the ACTA2-p.M49T variant, illustrating the marked elastic tissue fragmentation and adventitial fibrosis, all without any significant inflammatory infiltration. There is also significantly increased smooth muscle cellularity within the media, with only occasional Ki-67-positive cells (arrows), indicating modest proliferative activity. The second column illustrates similar images from a man in his sixties with hypertensive aortopathy following surgical resection of a proximal aortic aneurysm. Elastic tissue fragmentation is substantially diminished, with minimal adventitial fibrosis, relatively normal medial cellularity, and rare Ki-67-positive cells.

## Discussion

In summary, we utilized a new single-nucleus isolation and RNA-sequencing method for fixed pathology specimens to characterize aortic cellular heterogeneity in a patient harboring an ACTA2 variant of uncertain significance (p.M49T). Although this specific mutation lacks established pathogenicity in clinical databases, the early age of onset, positive family history and size of the aneurysm all suggest a genetic cause for TAA. Our single-cell data reveal expansion of VSMCs within the ascending thoracic aorta, accompanied by widespread alterations in VSMC gene expression in the ACTA 2-p.M49T sample. RNA-trajectory analysis implicated a pro-proliferative phenotypic transition among VSMCs in the ACTA2-p.M49T aorta, providing additional insights into its pathogenesis. These findings support an emerging paradigm originally proposed by a recent study that α-smooth muscle actin (αSMA) serves as a nuclear regulator of gene expression, such that *ACTA2* mutations disrupt VSMC transcriptional programs to transition cells towards a proliferative phenotype.^3^ This new paradigm therefore extends the pathogenesis of *ACTA2*-associated TAA beyond the structural role of ACTA2 as an extracellular matrix protein.^4^ In this case, we offer preliminary insight into the VSMC cell state transition in *ACTA2*-associated disease to a new genetic variant seen in a young patient with TAA. Our analysis of aneurysmal aortic tissue from this patient highlights that multiple *ACTA2* missense mutations that span its functional domains promote VSMC proliferation and global transcriptional changes, which may contribute to the accelerated TAA manifestations seen in these patients.

In broader terms, these findings show the combined power of genetic and single cell RNA-sequencing analysis to determine the pathogenicity of a novel variant. This analysis employs a protocol for performing snRNA-seq on formalin-fixed paraffin-embedded (FFPE) tissue, which facilitates the extension of prior research on ACTA2 to other mutations associated TAA. Historically, such investigations have been impeded by limited access to clinical samples, leading to reliance on laborious mechanistic studies in immortalized cell lines or iPSC-derived model systems. Our protocol overcomes this limitation by enabling transcriptional and cell-type-specific analysis of stored clinical samples obtained from long-term storage. To enable comparisons with single cell data obtained from other patients with variants of uncertain significance we have shared our data (https://markpepin.shinyapps.io/ACTA2_pMet49Thr_snRNA_Seq/). Future studies can directly compare multi-omic genetic and cell-type specific gene expression data to determine the role of genetic variants in TAA pathogenesis.

## Online Methods

### ACTA2-p.Met49Thr Mutation

CTA2, Exon 3, c.146T>C (p.Met49Thr) is a heterozygous SNP (rs869025352) labelled a “likely pathogenic” variant that substitutes methionine, a neutral and non-polar amino acid, with threonine, a neutral and polar amino acid, at codon 49 of exon 3 within the gene encoding ACTA2 (p.Met49Thr). It has also been observed to segregate with disease in related individuals. ClinVar contains an entry for this variant (Variation ID: 519576). Advanced modeling of protein sequence and biophysical properties (such as structural, functional, and spatial information, amino acid conservation, physicochemical variation, residue mobility, and thermodynamic stability) performed at Invitae® indicates that this missense variant is not expected to disrupt ACTA2 protein function. This variant disrupts the p.Met49 amino acid residue in ACTA2. Other variant(s) that disrupt this residue have been observed in individuals with ACTA2-related conditions,^6^ which suggests that this may be a clinically significant amino acid residue. In summary, the currently available evidence indicates that the variant is pathogenic, but additional studies are needed to prove this conclusively.

### Sample selection and nuclear isolation

Formalin-fixed and paraffin-embedded (FFPE) sections of aortic tissue were obtained from the Brigham and Women’s Hospital Department of Pathology. For nuclei dissociation and isolation from FFPE, we adapted a protocol as previously described.^7^ Using 50µm FFPE scrolls, we dissociated nuclei using the Miltenyi Biotech FFPE Tissue Dissociation Kit (CG000632 RevA, 10X Genomics), which was then filtered through a 20-μm cell strainer to remove cellular debris, after which a small aliquot was stained with DAPI (4′,6-diamidino-2-phenylindole) staining solution and loaded onto a hemocytometer to confirm nuclear viability under a fluorescence microscope. Quality control of the extracted RNA was achieved using TapeStation analysis for RNA integrity.

### Single-nuclear RNA sequencing

10X Genomics platform was applied in accordance with manufacturer’s instructions through the Brigham and Women’s Center for Cellular Profiling, as previously published. Following isolation of nuclei from clinical samples of proximal aortic tissue that had been formalin-fixed and paraffin-embedded (FFPE), approximately 500k nuclei were hybridized to contain unique probe barcodes as specified (CG000527, 10X Genomics). From this, ∼20k nuclei were loaded onto a Chromium Q chip (PN-1000422, 10X Genomics) for single-nucleus RNA sequencing (snRNA-seq). Libraries were prepared and sequenced on an Illumina NovaSeq platform with paired-end dual-indexing (28 cycles for Read 1, 10 cycles for i7, 10 cycles for i5, and 90 cycles for Read 2). Demultiplexing was performed using *bcl2fastq* (v1.8.4), and .fastq files were processed with *CellRanger* (v7.0.1) using the GRCh38-2020-A reference genome. Raw *Cellranger* output data were filtered to remove of ambient RNA using *cellbender* in full running mode. The resulting ACTA2-p.Met49Thr (ACTA2-pM49T) / control (NG) datasets contained 17,938 / 14,055 cells, with a median of 1,008 / 1,853 unique molecular identifiers (UMIs) and 716 / 1,188 genes detected per cell and 69.78% / 75.67% reads confidently mapped, respectively. These output files, which constitute feature-barcode matrices and cluster assignments, were then imported into the R (4.5.0) environment using the *Seurat* package (5.0.1) for downstream preprocessing, annotation, and differential expression.

To improve data quality, cells with fewer than 200 detected genes, as well as those containing more than 10% mitochondrial gene content, were excluded from downstream analysis. Normalization and scaling were performed using the “LogNormalize” method with a scaling factor of 10,000. Highly variable genes were identified using the “vst” method, and the 10,000 genes with the highest variance were retained for subsequent analysis. Unsupervised clustering via Uniform Manifold Approximation and Projection (UMAP) was performed on the scaled and variable gene expression data, and the top 30 principal components were used to define Louvian clustering with the “FindClusters” function (resolution = 0.2). Uniform Manifold Approximation and Projection (UMAP) was employed for dimensionality reduction and visualization of the clustered data. Gene-set enrichment analysis for all comparisons was performed using the 2023 Human WikiPathway curated database within *EnrichR* (3.2). Trajectory analysis of vascular smooth muscle cells (“VSMC”) populations was performed using *Monocle3* (1.4.22) in the R computing environment, as previously described.^8^

## Data Availability

All data produced in the present study are available upon reasonable request to the authors

## Data availability

All coding scripts and supplementary figures generated in the current study are available as an online supplement through the GitHub data sharing repository: https://github.com/mepepin/snRNA_ACTA2.p.Met49Thr. Additionally, the processed data are made fully accessible using an interactive web-based interface at the following link: https://markpepin.shinyapps.io/ACTA2_pMet49Thr_snRNA_Seq/.

## Statistical analysis

For all pairwise comparisons, the Shapiro-Wilk test for normality was performed to determine the most appropriate statistical test. Statistical comparisons were achieved using two-tailed t-tests between DCM and CON in the cell size and ANP intensity as well as qPCR experiments. All data are reported as mean ± standard deviation unless otherwise specified.

## Acknowledgements

We thank the Brigham and Women’s sequencing core facility for their assistance with processing the samples.

## Sources of funding

This work was funded by National Institutes of Health Grants U01-HL166060, R01-HL164811, DP2-HL152423, and the Eugene Braunwald Junior Faculty Scholar Award (R.M.G.). M.E.P. received funding support for sequencing through a junior investigator award from the Brigham and Womens’ Hospital Department of Medicine.

## Disclosures

RMG received research funding from Pfizer unrelated to this project.

## Authors’ contributions

M.E.P. performed all bioinformatics analysis, created figures, and wrote the manuscript with R.M.G. R.M. performed histologic sectioning and analysis. R.M.G. is the guarantors of this work and accepts responsibility for its integrity. All authors read, edited, and approved the final manuscript.

## Notes

### Competing Interest Statement

The authors have declared no competing interest.

### Author Declarations

This study was approved by the Institutional Review Board of Brigham and Womens Hospital (IRB protocol #2021P001077) and conducted in accordance with the Declaration of Helsinki. The requirement for written informed consent was waived due to the use of anonymized data.

